## Supplemental Tables and Figures for "Rapid Biphasic Decay of Intact and Defective HIV DNA Reservoir During Acute Treated HIV Disease"

**Supplementary Fig. 1: The UCSF Treat Acute HIV cohort study participants**. A total of 67 participants met inclusion criteria for acute HIV, defined as <100 days since the estimated date of detected HIV infection (EDDI) using the Infection Dating Tool (<https://tools.incidence-estimation.org/idt/>). The numbers of study participants by Fiebig stages (I-V) of HIV recency are also shown (https://doi.org:10.1097/01.aids.0000076308.76477.b8). PrEP = Pre-exposure prophylaxis with tenofovir disoproxil fumarate/emtricitabine (TDF/FTC).

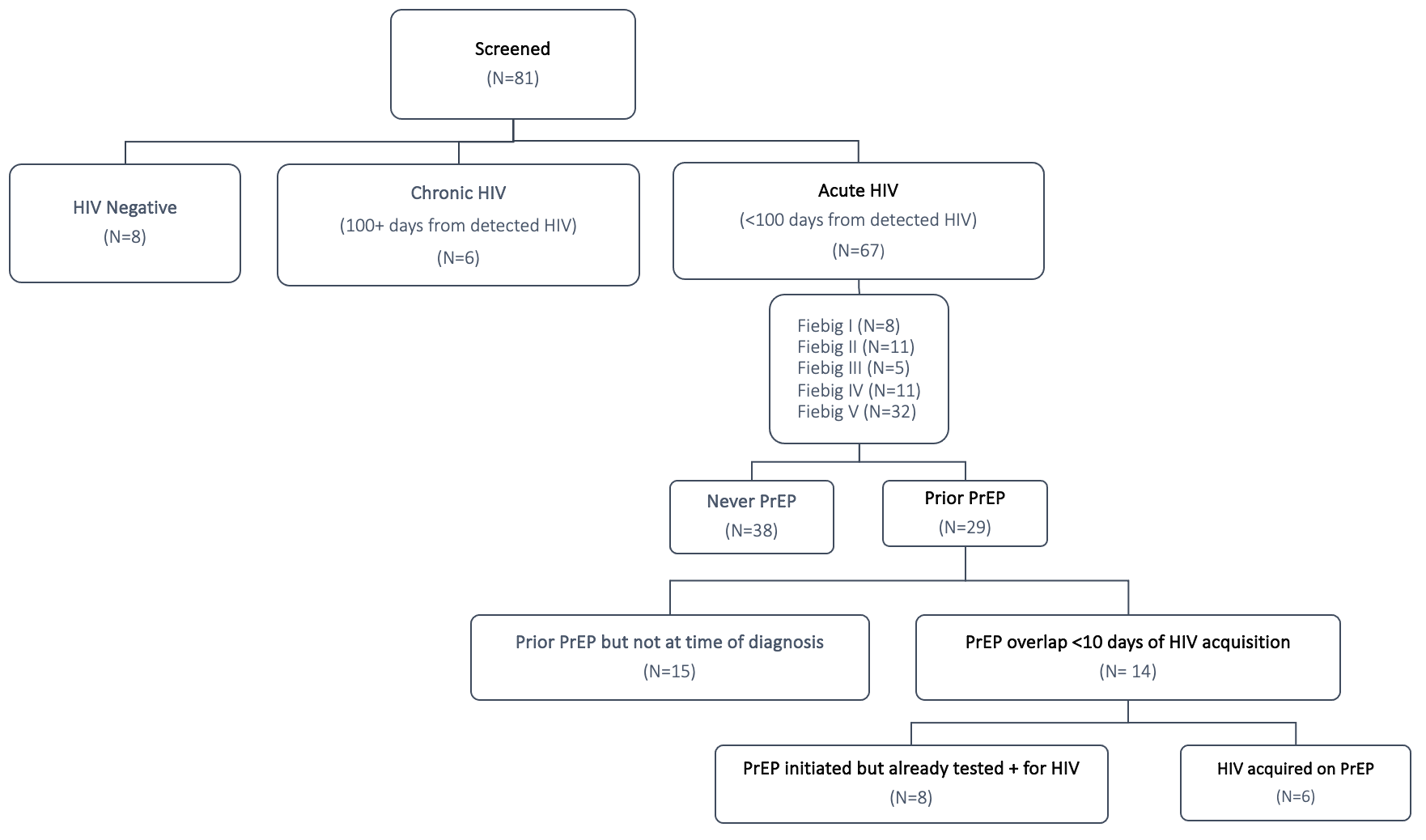

**Supplementary Fig. 2: Calculation of estimated dates of detected HIV infection**. The estimated dates of detected HIV infection (EDDI), along with a “confidence interval” for early probable (EP-EDDI) and late probable (LP-EDDI) dates, were calculated using participants’ clinical test results as well as baseline study visit confirmatory assay results.

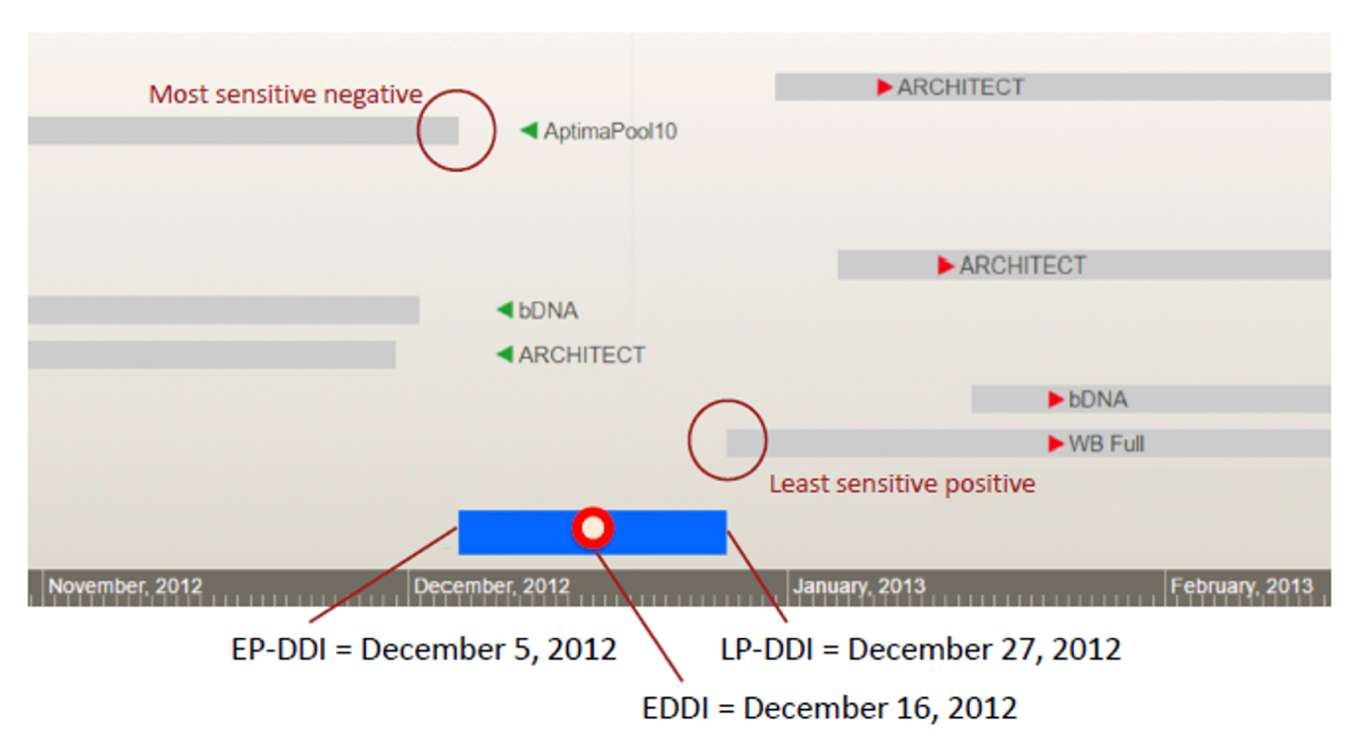

**Supplementary Fig. 3: HIV-1/2 test results for study participants.** The proportion of study participants with either negative and/or indeterminate test results for HIV-1/2 p24 antigen/antibody assay (Architect) (a) and HIV-1/2 differentiation (Geenius) antibody assay (b) at baseline study visit were consistent with rates with our San Francisco Department of Public Health rates (27% and 28%, respectively).

**a. b.**

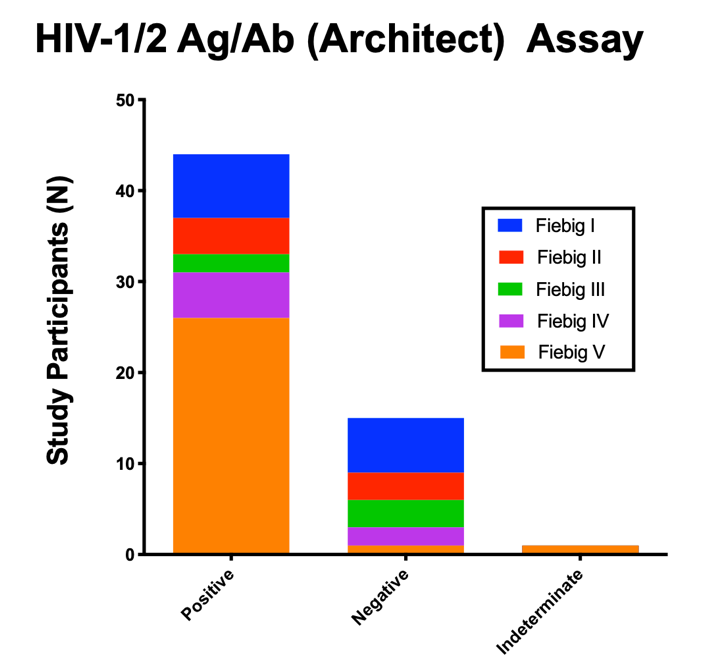

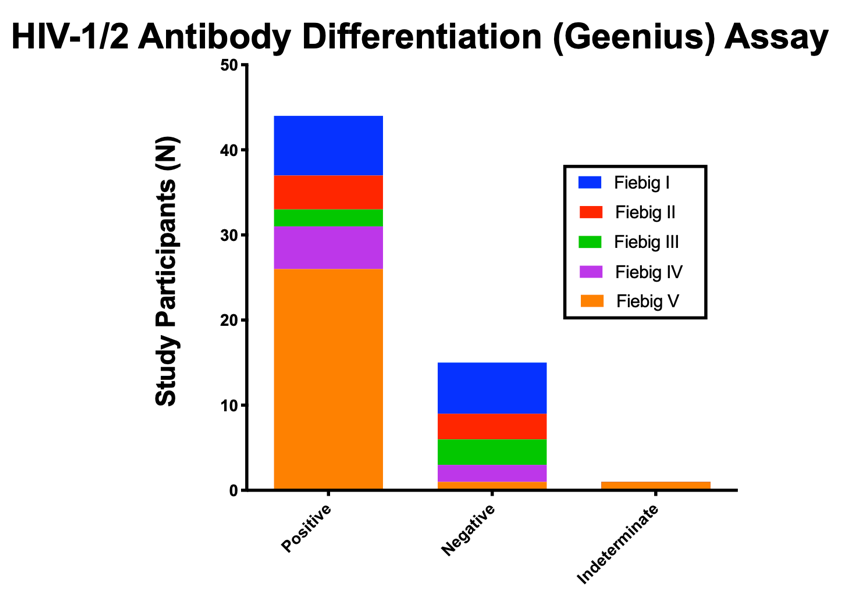

**Supplementary Fig. 4:** **HIV intact and defective DNA decay patterns by self-reported gender and prior PrEP use.** Observed HIV intact and defective DNA data, highlighting the one cisgender female (yellow line) and one transgender female (blue line) participants (a). Participants reporting PrEP use within 10 days of HIV diagnosis fell into two categories: 6 participants who acquired HIV while already taking PrEP (yellow lines), and 8 participants who were found to already have acquired HIV at the time of PrEP initiation (blue lines). All other study participants are shown as grey lines.

**a.**

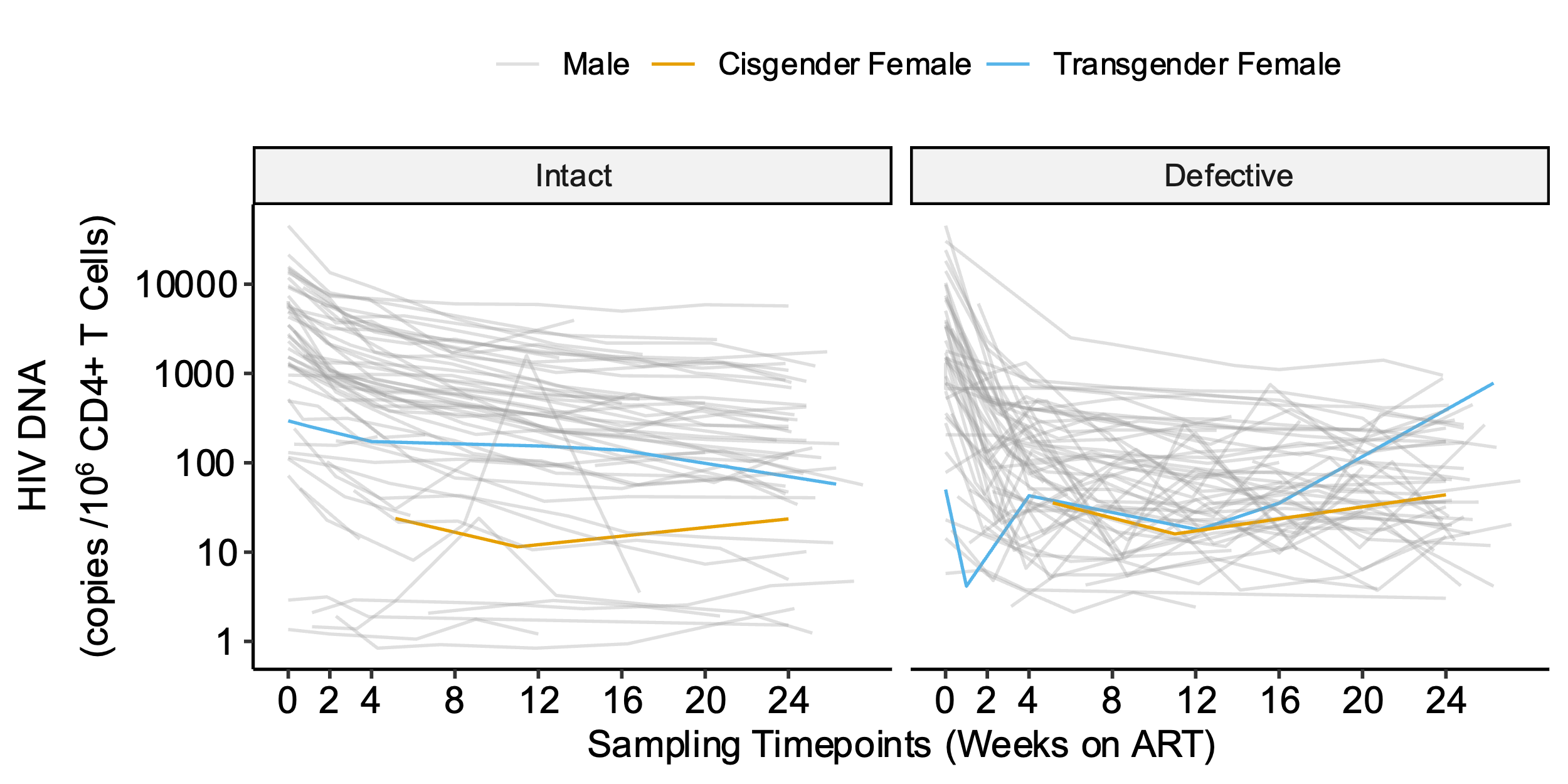

**b.**

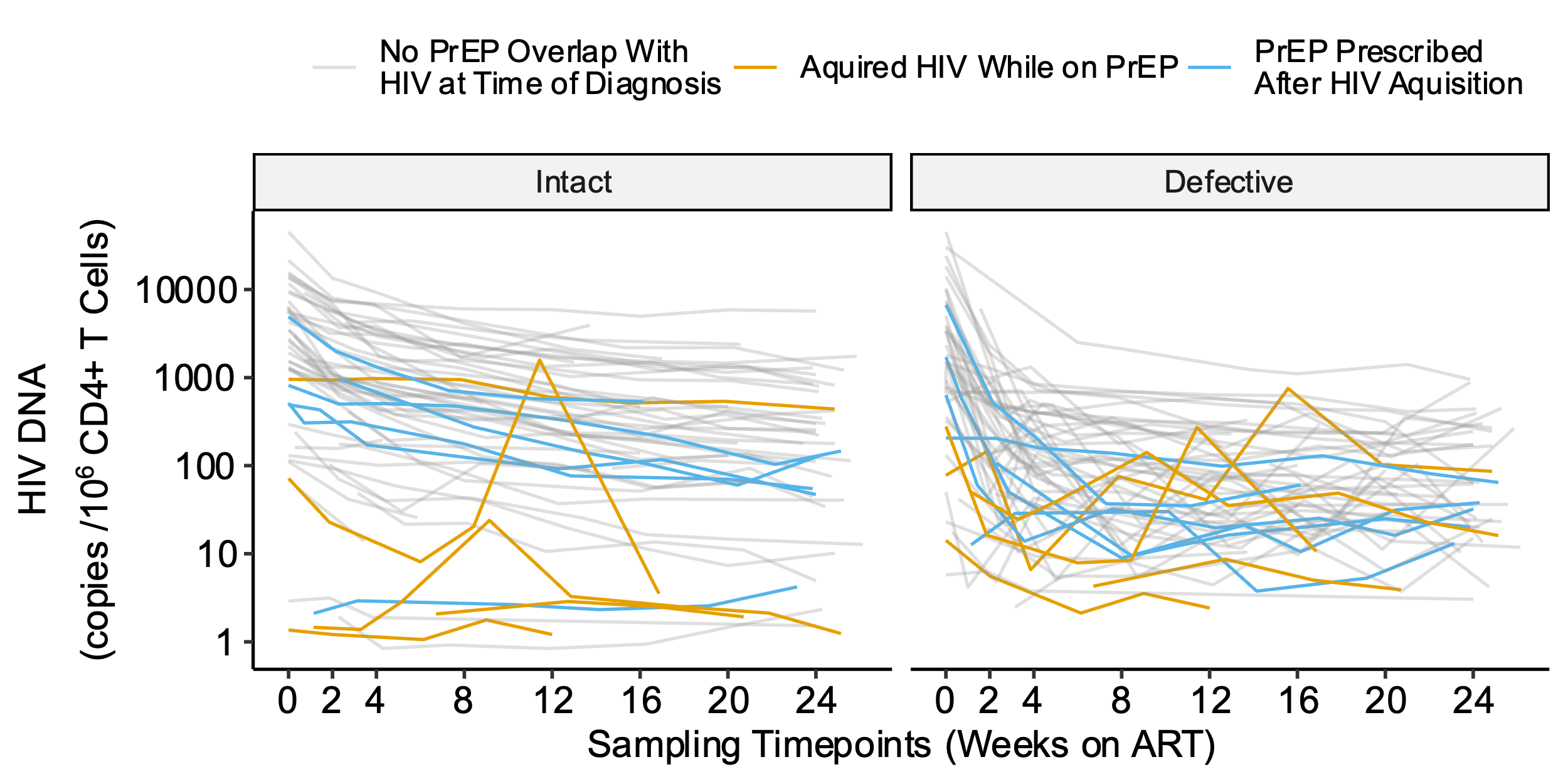

**Supplementary Fig. 5: Fine tuning of biphasic decay model inflection points from weeks 0-52.** A total of 65.7% of the study participants continued in follow-up beyond 24 weeks. We further refined our estimates for the inflection point, τ, by testing sequential half-week windows from 0 to 52 weeks and comparing the minimum prediction error using the leave-one-out mean absolute error (MAE, upper panels) or the leave-one-out mean squared error (MSE, lower panels). An inflection point of τ = 5 weeks (vertical dashed line) remained the best fit decay pattern for both HIV intact (left panels) and defective (right panels) DNA out to 52 weeks of ART. Red dots denote the best τ for each model and prediction error metric.

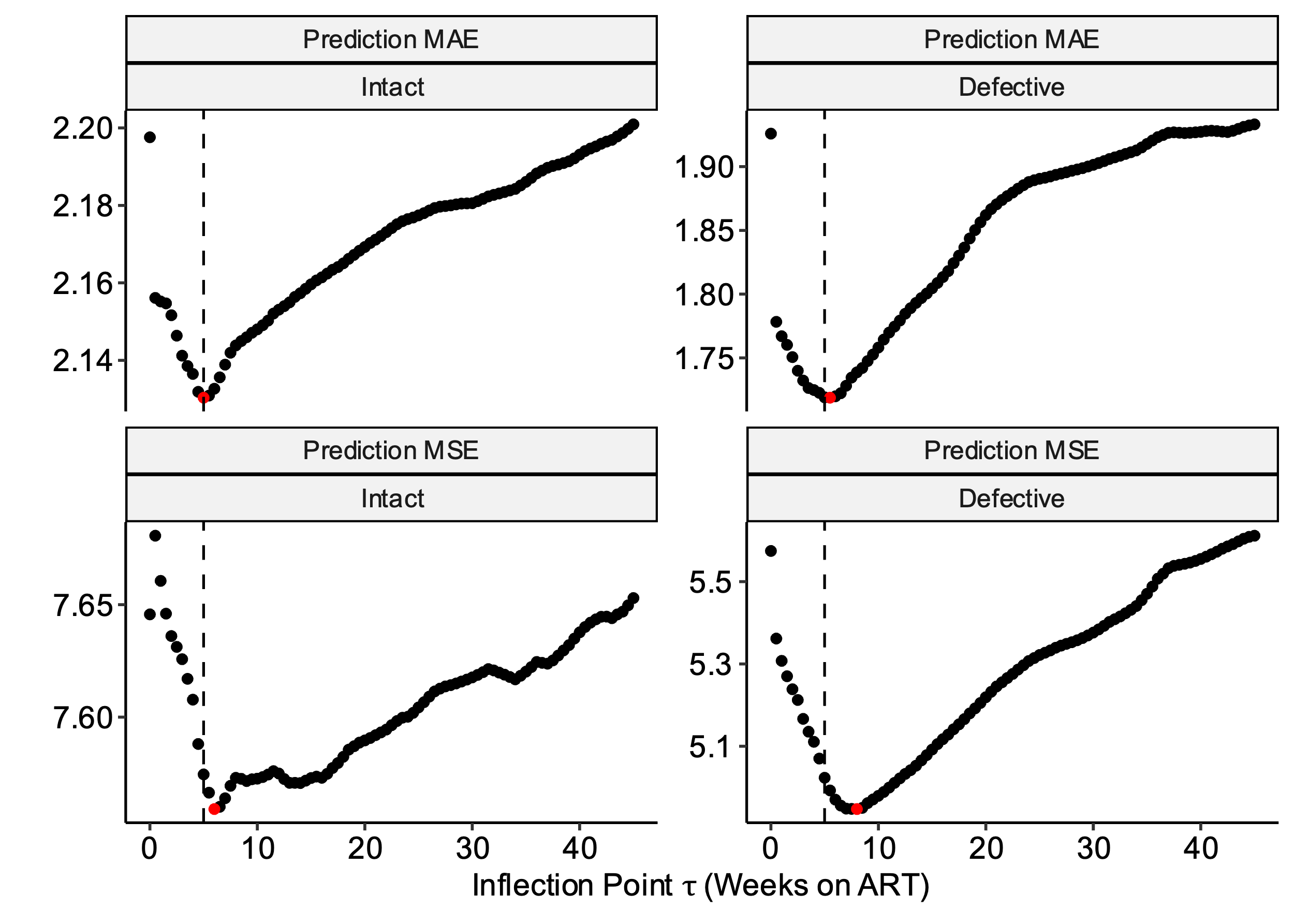

**Supplementary Table 1.** **Model estimates for HIV intact and defective DNA reservoir decay rates during acute treated HIV**. Slope and half-life (t_1/2_) estimates of HIV intact and defective decay rates for unadjusted (a-b) and adjusted models (b-c) during frequently sampled timepoints (weeks 0-24; a, c) and extended out to one year (weeks 0-52; b, d). Adjusted models included covariates for initial CD4+ T cell count, pre-ART HIV RNA, and timing of ART initiation.

**a. Unadjusted models (0-24 weeks)**

| **(N=61 participants)** | **Slope Estimate** | **Slope SE** | **P value** | **Decay Rate per Week (%)** | **Half Life (Week)** | **Half Life (lower 95% CI)** | **Half Life**  **(upper 95% CI)** |
| --- | --- | --- | --- | --- | --- | --- | --- |
| **Phase 1: 0 to 5 weeks** |  |  |  |  |  |  |  |
| **HIV Intact DNA** | -0.357 | 0.0297 | 2.03e-27 | 21.9 | 2.80 | 2.35 | 3.26 |
| **HIV Defective DNA** | -0.745 | 0.0549 | 3.28e-33 | 40.3 | 1.34 | 1.15 | 1.54 |
| **Phase 2: 5 to 24 weeks** |  |  |  |  |  |  |  |
| **HIV Intact DNA** | -0.0692 | 0.00784 | 8.87e-17 | 4.68 | 14.5 | 11.2 | 17.7 |
| **HIV Defective DNA** | 0.00184 | 0.0145 | 8.99e-01 | -0.127 | -544 | -8960 | 7880 |

**b. Unadjusted models (0-52 weeks)**

| **(N=61 participants)** | **Slope Estimate** | **Slope SE** | **P value** | **Decay Rate per Week (%)** | **Half Life (Week)** | **Half Life (lower 95% CI)** | **Half Life**  **(upper 95% CI)** |
| --- | --- | --- | --- | --- | --- | --- | --- |
| **Phase 1: 0 to 5 weeks** |  |  |  |  |  |  |  |
| **HIV Intact DNA** | -0.384 | 0.0280 | 3.22e-34 | 23.4 | 2.61 | 2.23 | 2.980 |
| **HIV Defective DNA** | -0.751 | 0.0512 | 4.55e-38 | 40.6 | 1.33 | 1.15 | 1.51 |
| **Phase 2: 5 to 52 weeks** |  |  |  |  |  |  |  |
| **HIV Intact DNA** | -0.0539 | 0.00498 | 1.30e-23 | 3.67 | 18.6 | 15.2 | 21.9 |
| **HIV Defective DNA** | 0.0102 | 0.00910 | 2.62e-01 | -0.711 | -97.9 | -269 | 73.0 |

**c. Adjusted models (0-24 weeks)**

| **(N=61 participants)** | **Slope Estimate** | **Slope SE** | **P value** | **Decay Rate per Week (%)** | **Half Life (Week)** | **Half Life (lower 95% CI)** | **Half Life**  **(upper 95% CI)** |
| --- | --- | --- | --- | --- | --- | --- | --- |
| **Phase 1: 0 to 5 weeks** |  |  |  |  |  |  |  |
| **HIV Intact DNA** | -0.354 | 0.0280 | 1.08e-29 | 21.7 | 2.83 | 2.39 | 3.27 |
| **HIV Defective DNA** | -0.735 | 0.0516 | 8.26e-36 | 39.9 | 1.36 | 1.17 | 1.55 |
| **Phase 2: 5 to 24 weeks** |  |  |  |  |  |  |  |
| **HIV Intact DNA** | -0.0648 | 0.00742 | 1.63e-16 | 4.39 | 15.4 | 12.0 | 18.9 |
| **HIV Defective DNA** | 0.0066 | 0.0137 | 6.32e-01 | -0.456 | -152 | -777 | 472 |

**d. Adjusted models (0-52 weeks)**

| **(N=61 participants)** | **Slope Estimate** | **Slope SE** | **P value** | **Decay Rate per Week (%)** | **Half Life (Week)** | **Half Life (lower 95% CI)** | **Half Life**  **(upper 95% CI)** |
| --- | --- | --- | --- | --- | --- | --- | --- |
| **Phase 1: 0 to 5 weeks** |  |  |  |  |  |  |  |
| **HIV Intact DNA** | -0.377 | 0.0264 | 2.00e-36 | 23.0 | 2.65 | 2.29 | 3.02 |
| **HIV Defective DNA** | -0.737 | 0.0481 | 1.05e-40 | 40.0 | 1.36 | 1.18 | 1.53 |
| **Phase 2: 5 to 52 weeks** |  |  |  |  |  |  |  |
| **HIV Intact DNA** | -0.0517 | 0.00470 | 3.06e-24 | 3.52 | 19.3 | 15.9 | 22.8 |
| **HIV Defective DNA** | 0.0135 | 0.00856 | 1.17e-01 | -0.937 | -74.3 | -167 | 18.4 |

**Supplementary Fig. 6:** **Predicted decay patterns of HIV intact and defective DNA during acute treated HIV from weeks 0-52.** A total of 65.7% of the study participants continued in follow-up beyond 24 weeks. Decay patterns for observed (thin grey lines) HIV intact (left panel) and total defective (right panel) DNA closely fit with average model predictions (thick black lines). Sampling timepoints are labeled on the x-axis (including a week 2 study visit during which confirmatory HIV test results were disclosed). Average predicted participant predictions were made by taking the mean of $E_{i}$ (estimated time between HIV infection and ART initiation), $C_{i}$ (initial CD4+ T cell count), and $V_{i}$ (log_10_ pre-ART plasma viral load) across participants from final models.

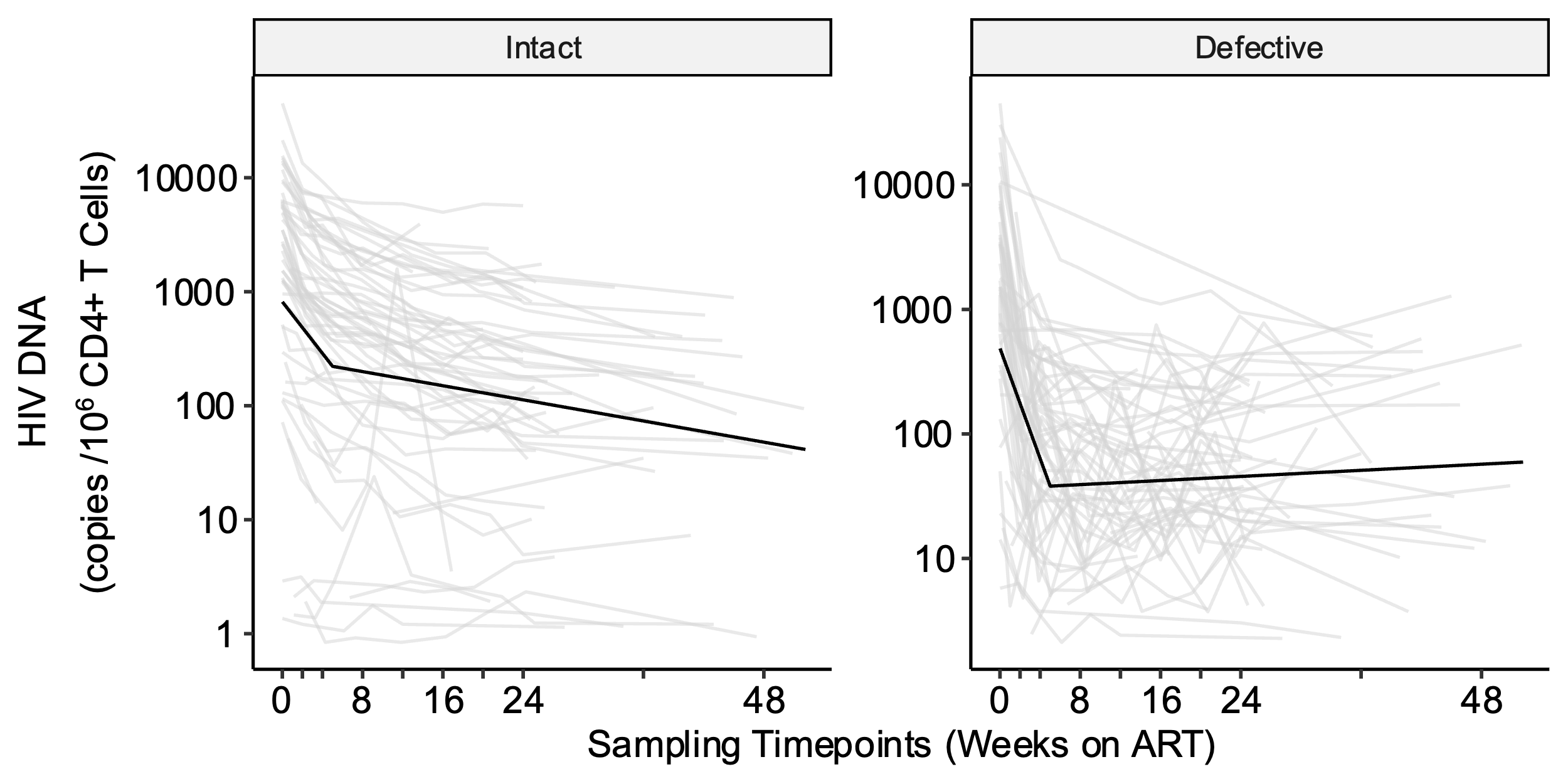

**Supplementary Fig. 7: Predicted versus observed plots show good model performance for both HIV intact and defective DNA**. Validation for the final models for intact and defective HIV DNA decay was initially performed by looking at the plots of predicted vs observed HIV DNA counts. These plots show that both models produce relatively unbiased estimates across the observed range of HIV DNA counts and that the residual variance in the defective reservoir is much higher than the intact reservoir. Red dashed line denotes the idealized fit where predicted values exactly equal observed values.

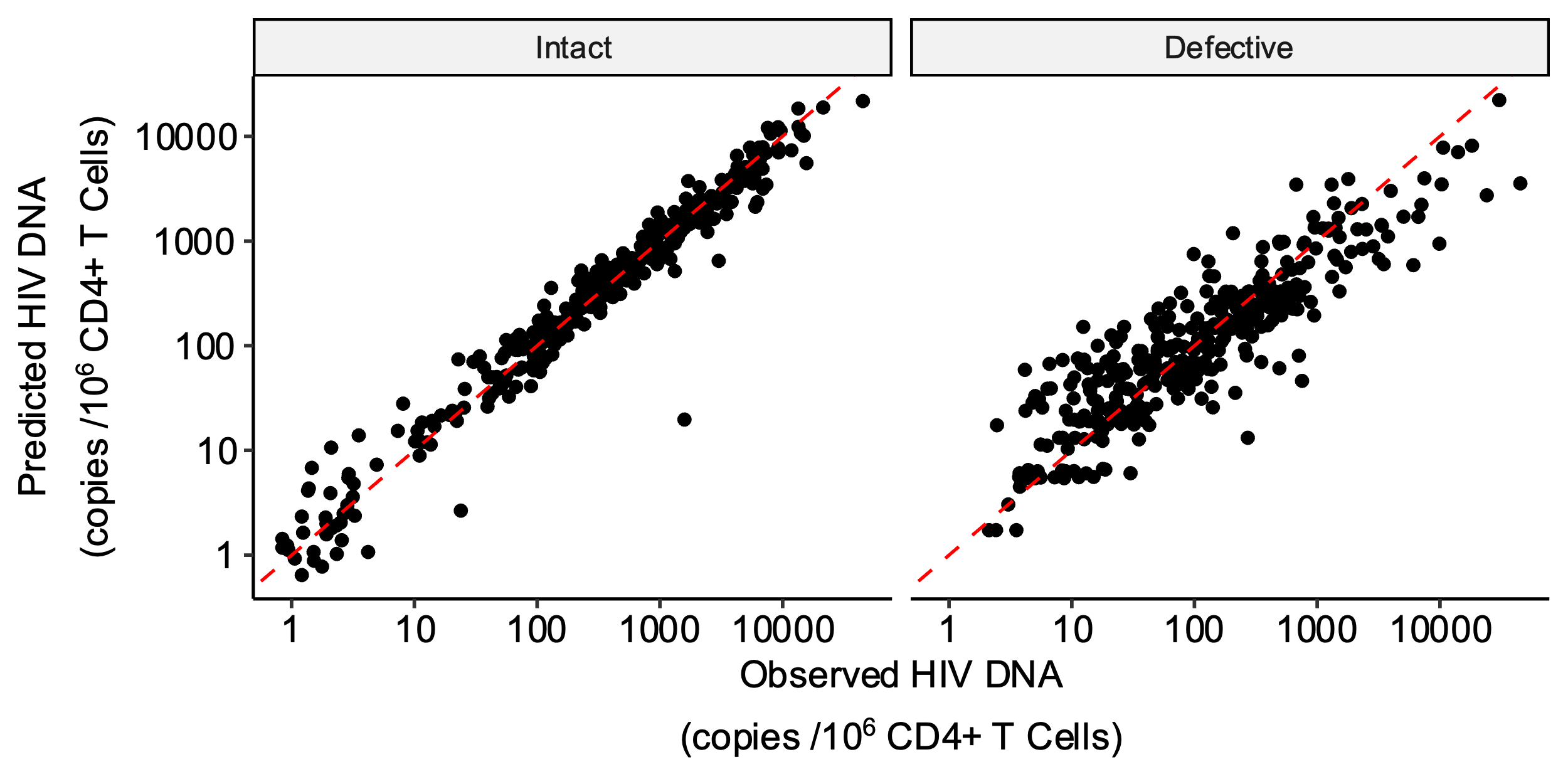

**Supplementary Fig. 8: Sensitivity analyses estimating HIV intact and defective DNA decay rates after excluding potential outlier clinical subgroups**. The final model (τ = 5 weeks) was fit on three clinically interesting sub-populations to assess if the influence of potential outlier data. Separate models were fit that excluded (a) participants reporting prior PrEP use (<10 days overlap between last PrEP use and estimated date of detected HIV infection), (b) participants with plasma viral load “blips” (defined as a one-time viral load >1000 copies/mL or two consecutive viral loads >100 copies/mL between weeks 0-24), and (c) participants with sudden increases in HIV intact DNA (defined as >50% increase between two consecutive measurements of HIV intact DNA during weeks 0-24). Models were fit using the cohort data (grey lines), but not the potential outlier data (red lines). The resulting predicted average participant HIV reservoir decay patterns are shown as thick black lines. Refer to **Supplementary Table 1** to get the sample sizes and half-life estimates for each sensitivity analysis.

**a.**

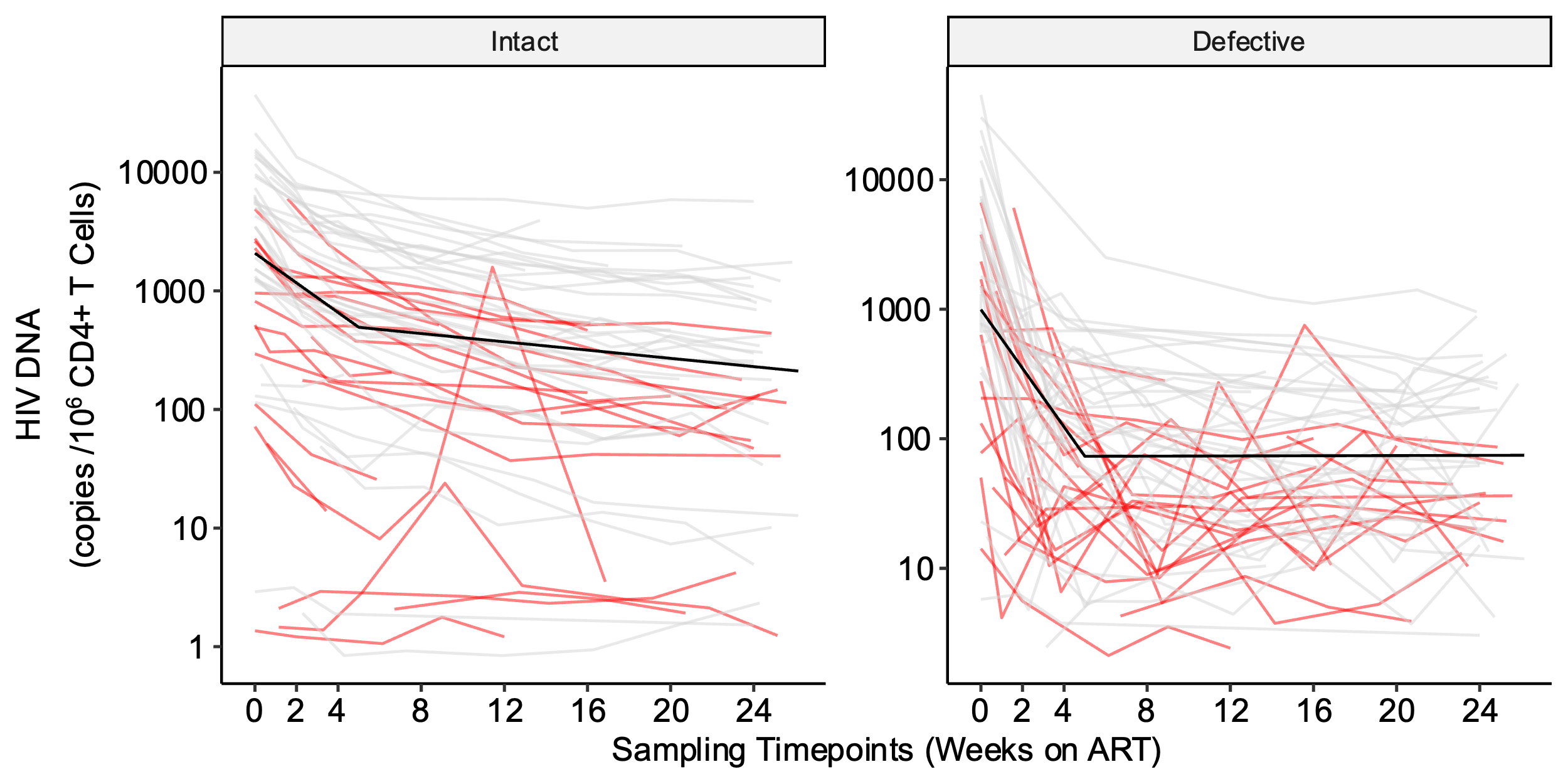

**b.**

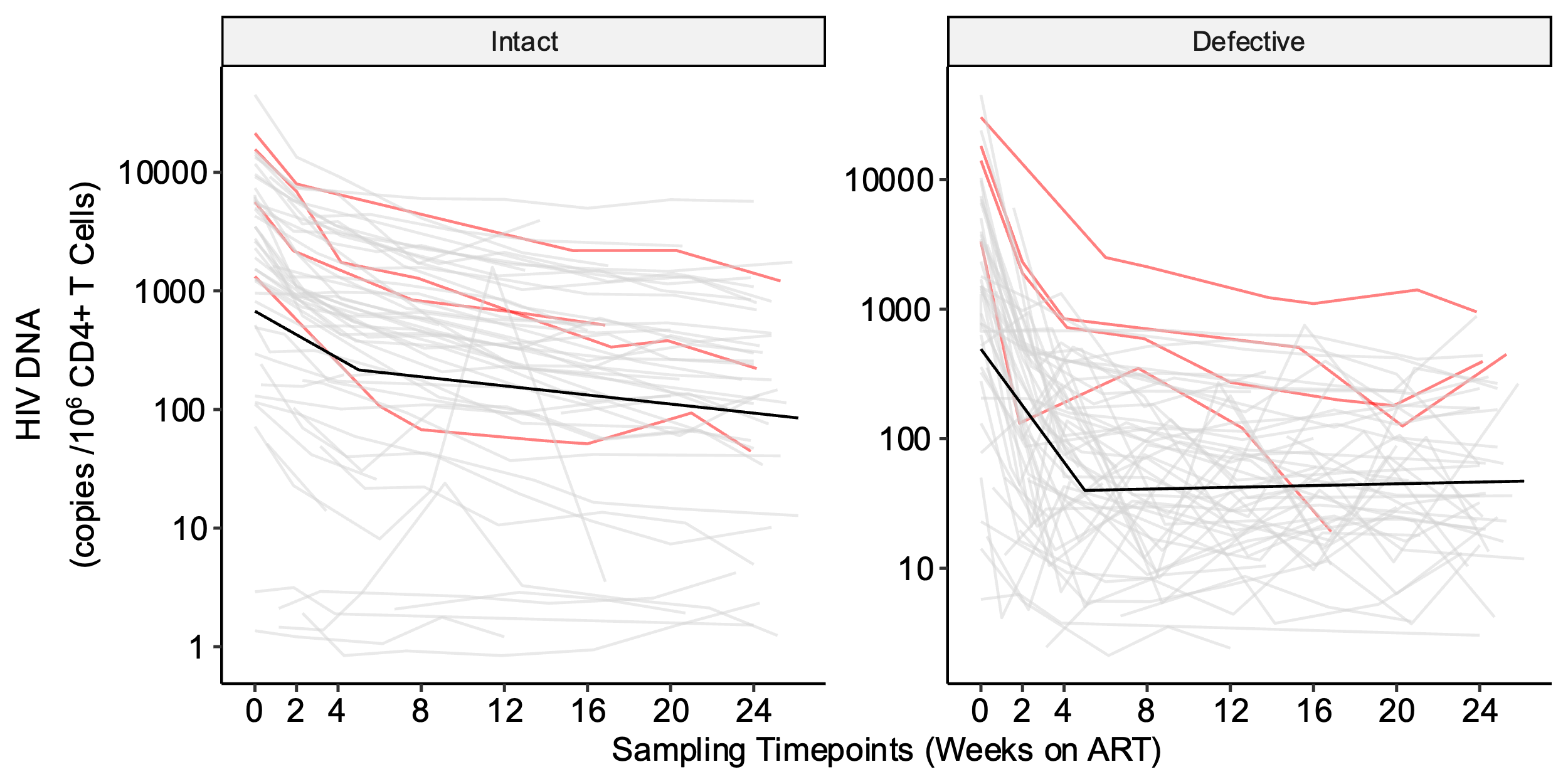

**c.**

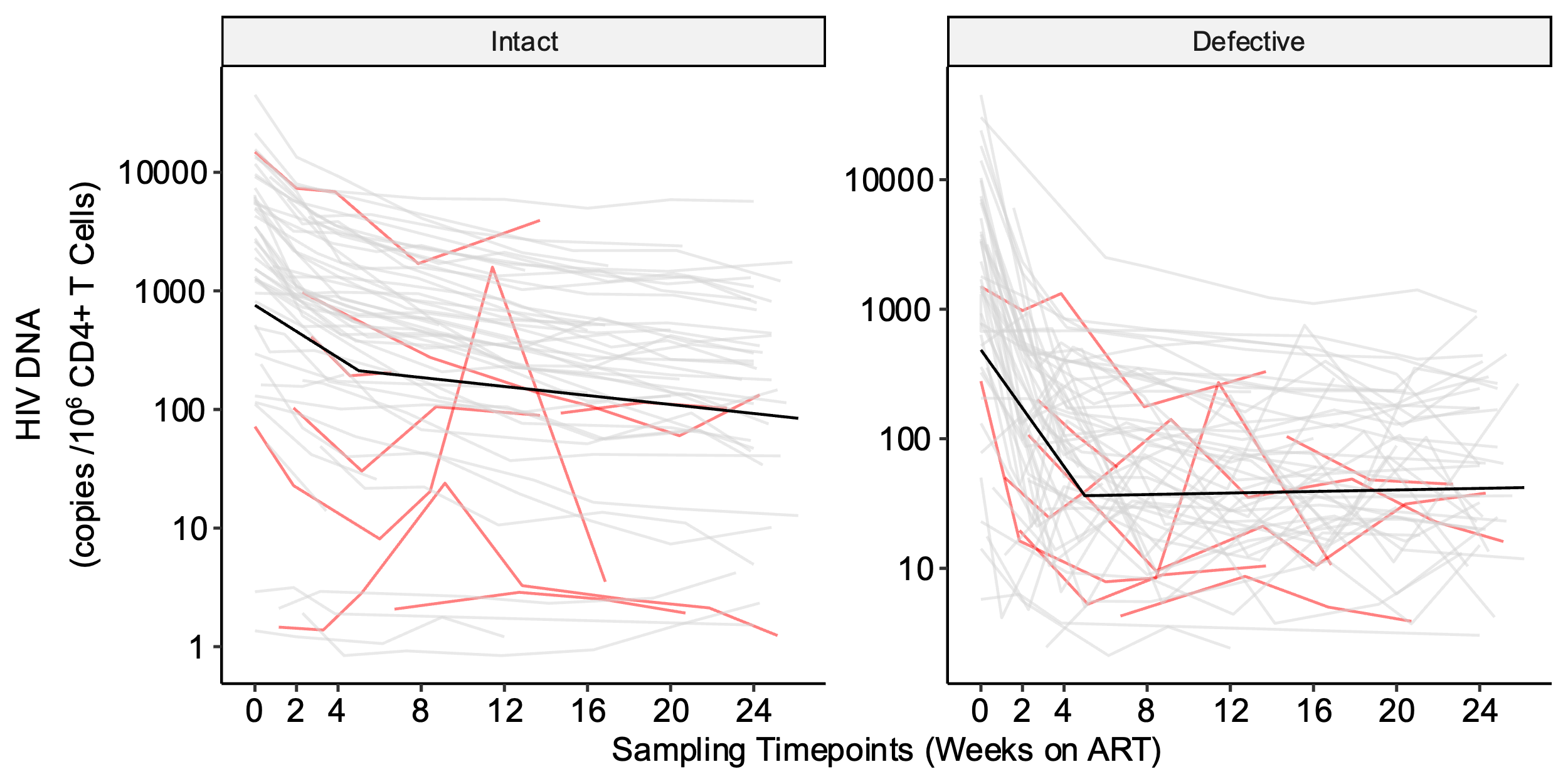

**Supplementary Fig. 9: Inflection point sensitivity analyses demonstrate some variability when excluding populations of potential outliers.** To test whether the final model inflection point selection of τ = 5 weeks was influenced by potential outlier data we performed τ estimation on three clinically interesting sub-populations. Separate models were fit that excluded (a) participants reporting prior PrEP use (<10 days overlap between last PrEP use and estimated date of detected HIV infection), (b) participants with plasma viral load “blips” (defined as a one-time viral load >1000 copies/mL or two consecutive viral loads >100 copies/mL between weeks 0-24), and (c) participants with sudden increases in HIV intact DNA (defined as >50% increase between two consecutive measurements of HIV intact DNA during weeks 0-24). A regular grid of possible τ was used (0-26 weeks by half-week) and the leave-one-out cross-validation (LOOCV) mean absolute prediction error (MAE) was computed for each candidate τ. Red dots denote the best τ for each model and prediction error metric and our selected inflection point (τ = 5) is shown with a dashed vertical line. Refer to **Supplementary Table 1** to get the sample size for each sensitivity analysis and to **Supplementary Fig. 9** to see which patients are excluded from each sensitivity analysis.

**a.**

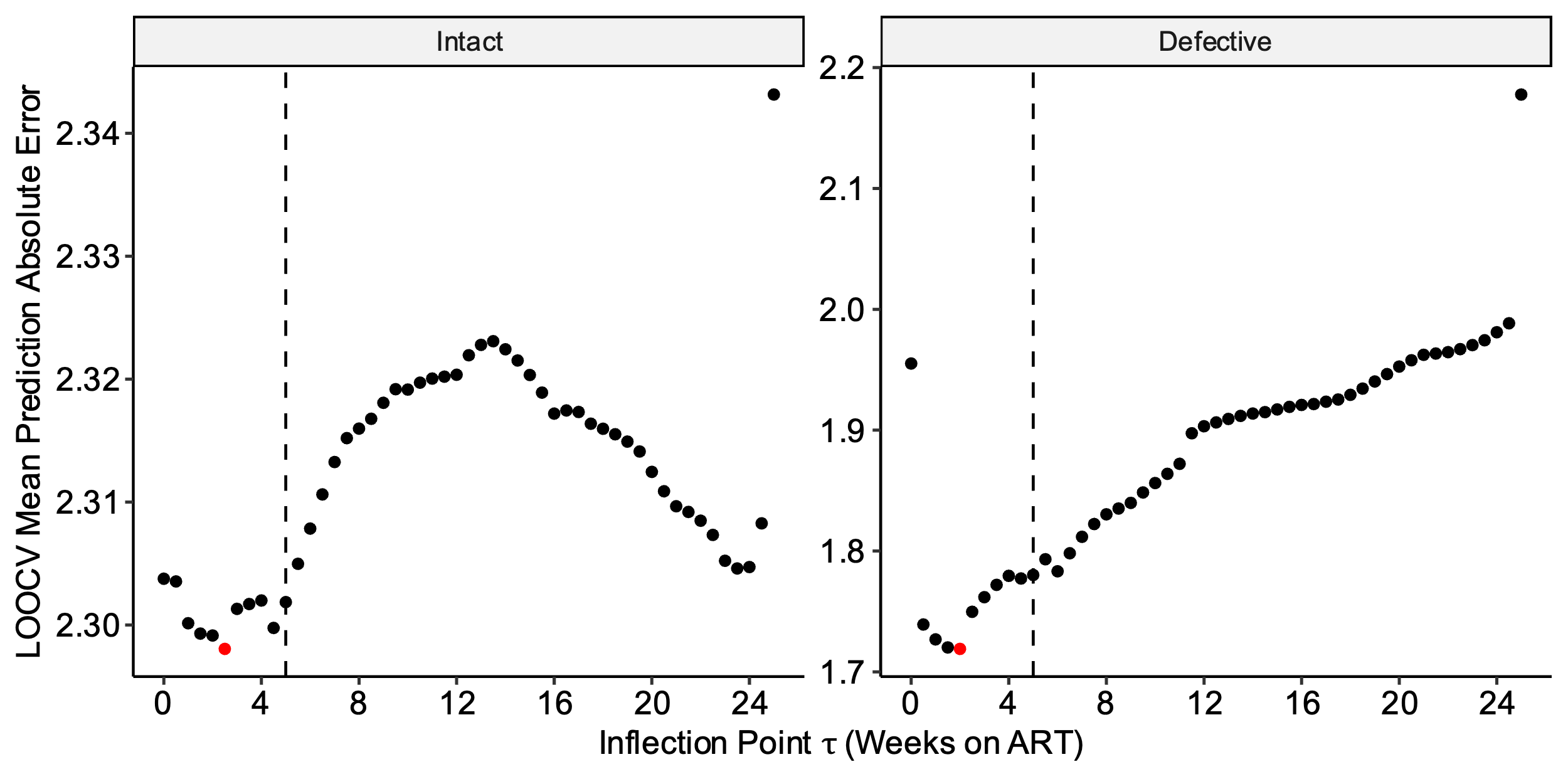

**b.**

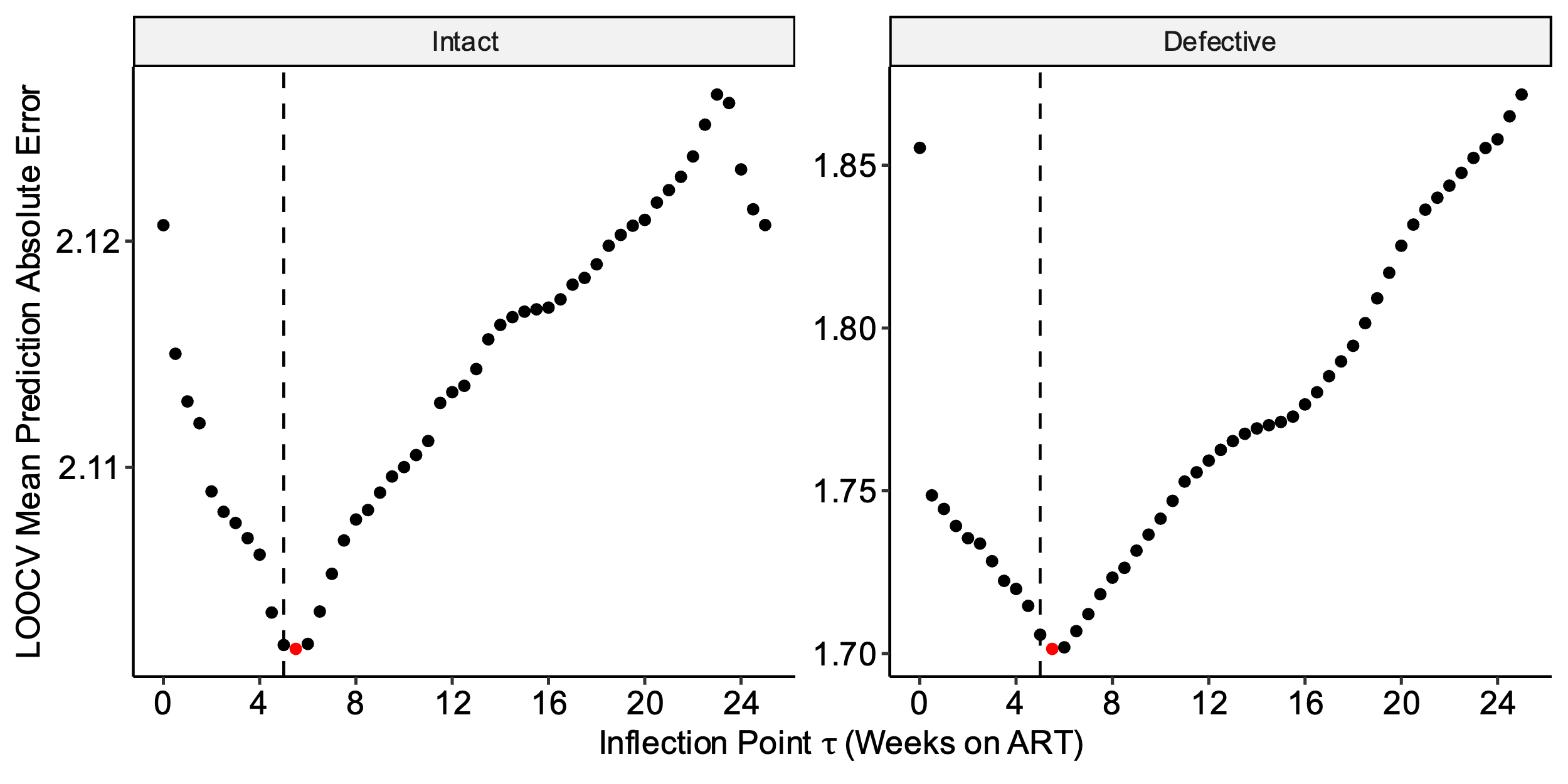

**c.**

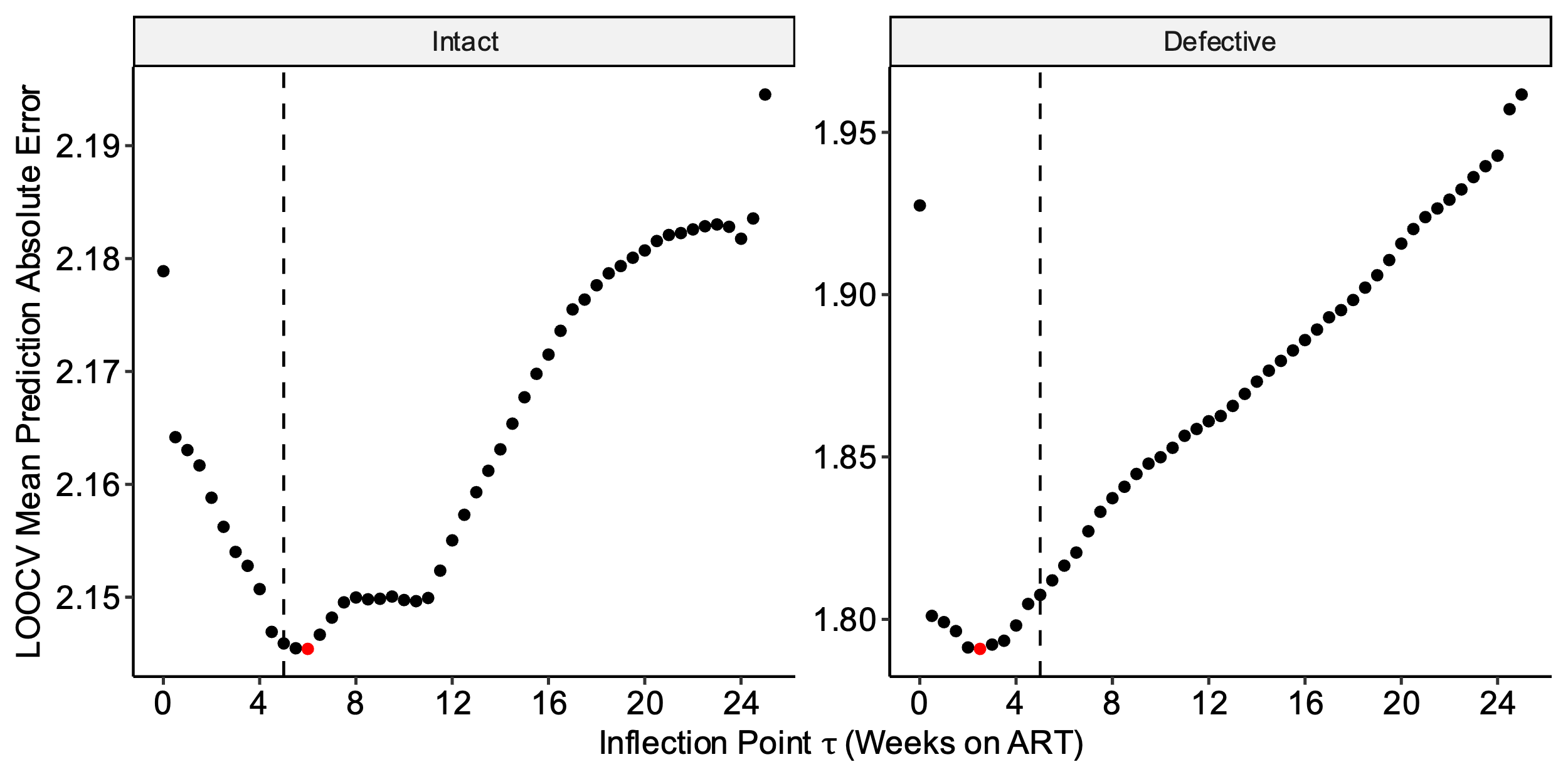

**Supplementary Table 2.** **Sensitivity analyses of HIV intact and defective DNA reservoir decay rates during acute treated HIV**. Slope and half-life estimates of HIV intact and defective decay rates after excluding participants reporting prior preexposure prophylaxis (PrEP) use within 10 days of estimated date of HIV infection (a), participants with plasma viral load “blips” (defined as a one-time viral load >1000 copies/mL or two consecutive viral loads >100 copies/mL between weeks 0-24) (b), and participants with sudden increases in HIV intact DNA (defined as >50% increase between two consecutive measurements of HIV intact DNA during weeks 0-24) (c). All models were adjusted for initial CD4+ T cell count, pre-ART HIV RNA, and timing of ART initiation.

**a. Excluding participants with recent PrEP use**

| **(N=37 participants)** | **Slope Estimate** | **Slope SE** | **P value** | **Decay Rate per Week (%)** | **Half Life (Week)** | **Half Life (lower 95% CI)** | **Half Life**  **(upper 95% CI)** |
| --- | --- | --- | --- | --- | --- | --- | --- |
| **Phase 1: 0 to 5 weeks** |  |  |  |  |  |  |  |
| **HIV Intact DNA** | -0.427 | 0.0247 | 1.37E-41 | 25.6 | 2.34 | 2.08 | 2.61 |
| **HIV Defective DNA** | -0.782 | 0.0610 | 6.02E-28 | 41.8 | 1.28 | 1.08 | 1.48 |
| **Phase 2: 5 to 24 weeks** |  |  |  |  |  |  |  |
| **HIV Intact DNA** | -0.0618 | 0.00639 | 1.99E-18 | 4.20 | 16.2 | 12.9 | 19.4 |
| **HIV Defective DNA** | 0.000852 | 0.0158 | 9.57E-01 | -0.0591 | -1170 | -43900 | 41500 |

**b. Excluding participants with viral blips**

| **(N=57 participants)** | **Slope Estimate** | **Slope SE** | **P value** | **Decay Rate per Week (%)** | **Half Life (Week)** | **Half Life (lower 95% CI)** | **Half Life**  **(upper 95% CI)** |
| --- | --- | --- | --- | --- | --- | --- | --- |
| **Phase 1: 0 to 5 weeks** |  |  |  |  |  |  |  |
| **HIV Intact DNA** | -0.333 | 0.0293 | 7.78E-25 | 20.6 | 3.00 | 2.48 | 3.52 |
| **HIV Defective DNA** | -0.737 | 0.0547 | 2.26E-32 | 40.0 | 1.36 | 1.16 | 1.56 |
| **Phase 2: 5 to 24 weeks** |  |  |  |  |  |  |  |
| **HIV Intact DNA** | -0.0652 | 0.00773 | 1.68E-15 | 4.42 | 15.3 | 118 | 18.9 |
| **HIV Defective DNA** | 0.0111 | 0.0145 | 4.45E-01 | -0.770 | -90.4 | -322 | 141 |

**c. Excluding participants with sudden increases in HIV intact DNA**

| **(N=54 participants)** | **Slope Estimate** | **Slope SE** | **P value** | **Decay Rate per Week (%)** | **Half Life (Week)** | **Half Life (lower 95% CI)** | **Half Life**  **(upper 95% CI)** |
| --- | --- | --- | --- | --- | --- | --- | --- |
| **Phase 1: 0 to 5 weeks** |  |  |  |  |  |  |  |
| **HIV Intact DNA** | -0.371 | 0.0225 | 8.85E-43 | 22.7 | 2.70 | 2.38 | 3.02 |
| **HIV Defective DNA** | -0.760 | 0.0545 | 8.25E-34 | 40.9 | 1.32 | 1.13 | 1.50 |
| **Phase 2: 5 to 24 weeks** |  |  |  |  |  |  |  |
| **HIV Intact DNA** | -0.0648 | 0.00603 | 1.05E-22 | 4.39 | 15.4 | 12.6 | 18.2 |
| **HIV Defective DNA** | 0.00915 | 0.0146 | 5.33E-01 | -0.636 | -109 | -452 | 233 |

**Supplementary Table 3.** **The effect of earlier ART initiation on HIV intact and defective DNA decay rates**. We performed bootstrapping to calculate the difference in half-life estimates for HIV intact and defective DNA, with each week that ART is initiated earlier. All models were adjusted for initial CD4+ T cell count and pre-ART HIV RNA.

|  | **N** | **Decrease in Half-Life per each Week Earlier ART Initiation** | **Decrease in Half-Life (lower 95% CI)** | **Decrease in Half-Life (upper 95% CI)** | **P value** |
| --- | --- | --- | --- | --- | --- |
| **Timing of ART** | 61 |  |  |  |  |
| **Phase 1: 0 to 5 weeks** |  |  |  |  |  |
| **HIV Intact DNA** |  | 0.0827 | 0.0203 | 0.145 | 9.40e-03 |
| **HIV Defective DNA** |  | 0.0579 | 0.0312 | 0.0846 | 2.12e-05 |
| **Phase 2: 5 to 24 weeks** |  |  |  |  |  |
| **HIV Intact DNA** |  | 1.08 | 0.316 | 1.84 | 5.55e-03 |
| **HIV Defective DNA** |  | 25.7 | -200 | 252 | 8.24e-01 |

**Supplementary Table 4.** **The effect of higher initial CD4+ T cell count or lower pre-ART HIV RNA on HIV intact and defective DNA decay rates**. We performed bootstrapping to calculate average half-life estimates for HIV intact and defective DNA for each quartile change in initial CD4+ T cell count or pre-ART HIV RNA. Models estimating the effect of initial CD4+ T cell count were adjusted for timing of ART initiation and pre-ART HIV RNA. Models estimating the effect of pre-ART HIV RNA were adjusted for timing of ART initiation and initial CD4+ T cell count.

|  | **N** | **Clinical Value** | **Estimated Reservoir Adjustment** | **SE** | **Estimate (lower CI)** | **Estimate (upper CI)** |
| --- | --- | --- | --- | --- | --- | --- |
| **Initial CD4+ Count** | 61 |  |  |  |  |  |
| HIV Intact DNA |  |  |  |  |  |  |
| 25^th^ percentile |  | 350 | 1.90 | 0.803 | 0.324 | 3.47 |
| 50^th^ percentile |  | 505 | -0.0498 | 0.0250 | -0.0988 | -0.000805 |
| 75^th^ percentile |  | 664 | -2.04 | 0.865 | -3.74 | -0.349 |
| HIV Defective DNA |  |  |  |  |  |  |
| 25^th^ percentile |  | 350 | 1.26 | 0.543 | 0.199 | 2.33 |
| 50^th^ percentile |  | 505 | -0.144 | 0.258 | -0.649 | 0.362 |
| 75^th^ percentile |  | 664 | -1.44 | 0.625 | -2.67 | -0.220 |
| **Log_10_ Pre-ART Viral Load** | 61 |  |  |  |  |  |
| HIV Intact DNA |  |  |  |  |  |  |
| 25^th^ percentile |  | 3.78 | -4.12 | 0.748 | -5.58 | -2.65 |
| 50^th^ percentile |  | 4.86 | 0.0987 | 0.0251 | 0.0495 | 0.148 |
| 75^th^ percentile |  | 5.67 | 3.27 | 0.594 | 2.10 | 4.43 |
| HIV Defective DNA |  |  |  |  |  |  |
| 25^th^ percentile |  | 3.78 | -3.71 | 0.878 | -5.43 | -1.99 |
| 50^th^ percentile |  | 4.86 | -0.762 | 0.571 | -1.88 | 0.356 |
| 75^th^ percentile |  | 5.67 | 2.15 | 0.572 | 1.03 | 3.27 |

**Supplementary Fig. 10:** **Fitted spline models to estimate the effect of higher initial CD4+ T cell count or lower pre-ART HIV RNA on HIV intact and defective DNA decay rates.** Fitted spline models (red lines) with corresponding 95% confidence intervals (blue dashed lines) are shown for HIV intact DNA (a) and HIV defective (b) DNA. Plots are shown for pre-ART HIV RNA (left panels) and initial CD4+ T cell count (right panels).

**a. HIV intact DNA**

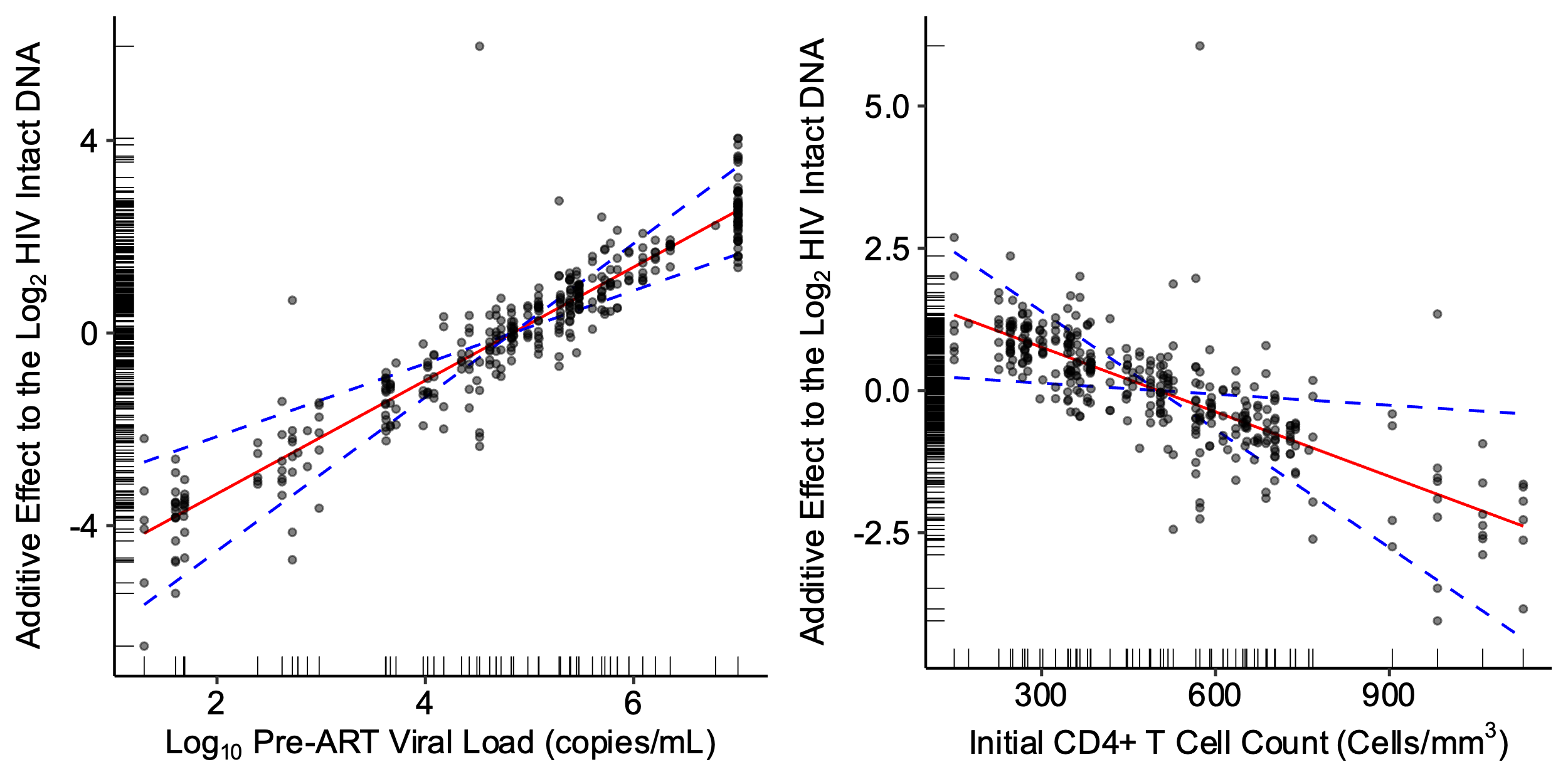

**b. HIV defective DNA**

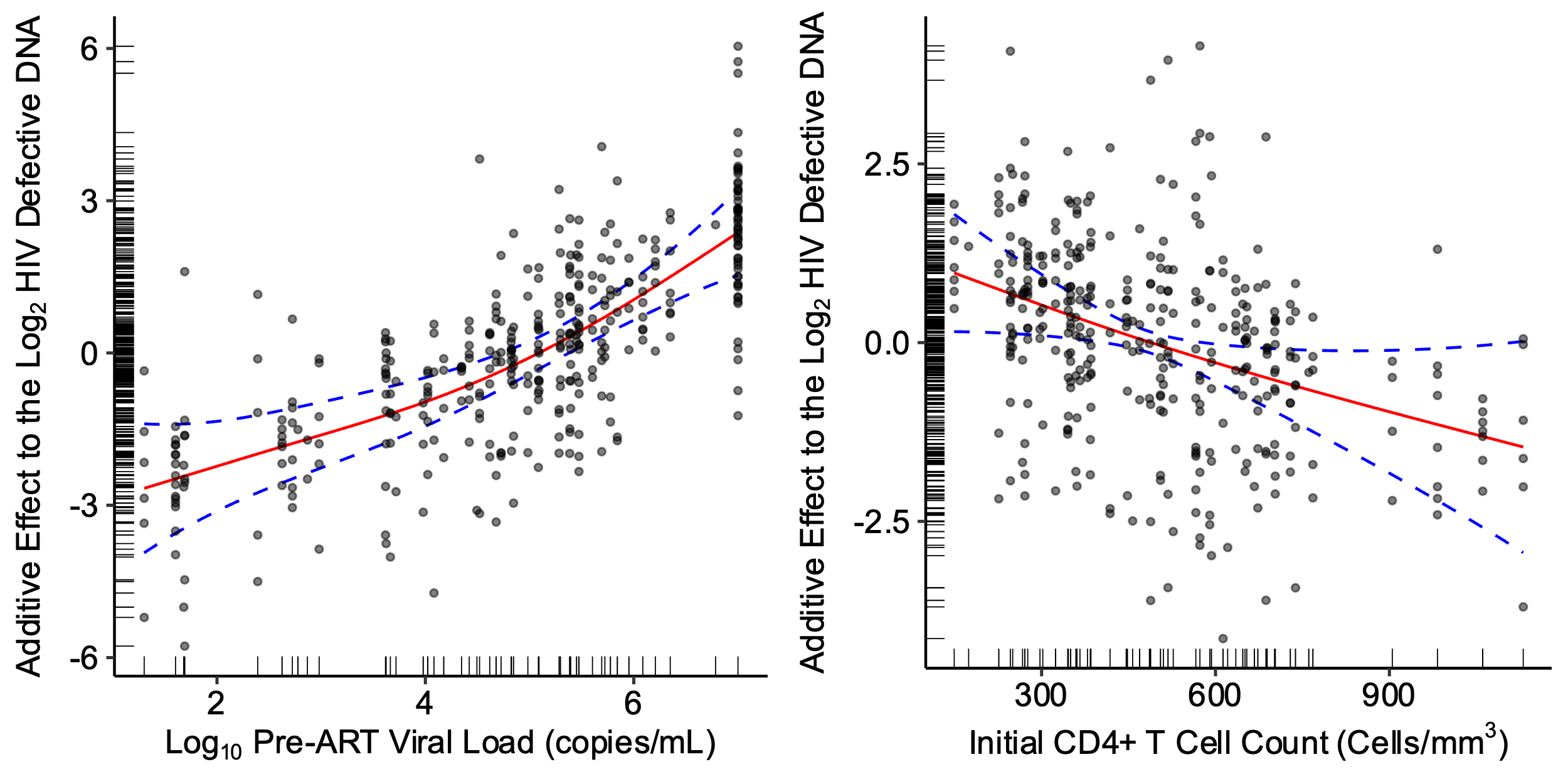

**Supplementary Fig. 11: Semiparametric monophasic, biphasic, and triphasic generalized additive models of plasma HIV RNA decay during weeks 0-24.** We performed bootstrapping to estimate the Akaike information criteria (AIC) value and 95% confidence intervals and compared monophasic, biphasic, and triphasic models for plasma HIV RNA (a). Using the triphasic model for plasma HIV RNA, we then determined the optimal inflection point(s), τ, by minimizing the predicted mean absolute error (MAE; top panel) using leave-one-out cross-validation or the predicted mean squared error (MSE; bottom panel) (b). Red dots denote the optimal inflection point(s), τ, for the model and loss metric. These analyses demonstrated an inflection point at 0.5 weeks (red dot at x-axis) and 4 weeks (red dot at y-axis), suggesting that a triphasic model best fit the data (b).

**a.**

| **HIV Reservoir** | **Monophasic (95% CI)** | **Biphasic (95% CI)** | **Triphasic (95% CI)** |
| --- | --- | --- | --- |
| **Plasma Viral Load** | 2408 (2288, 2503) | 1928 (1808, 2026) | 1908 (1747, 2037) |

**b.**

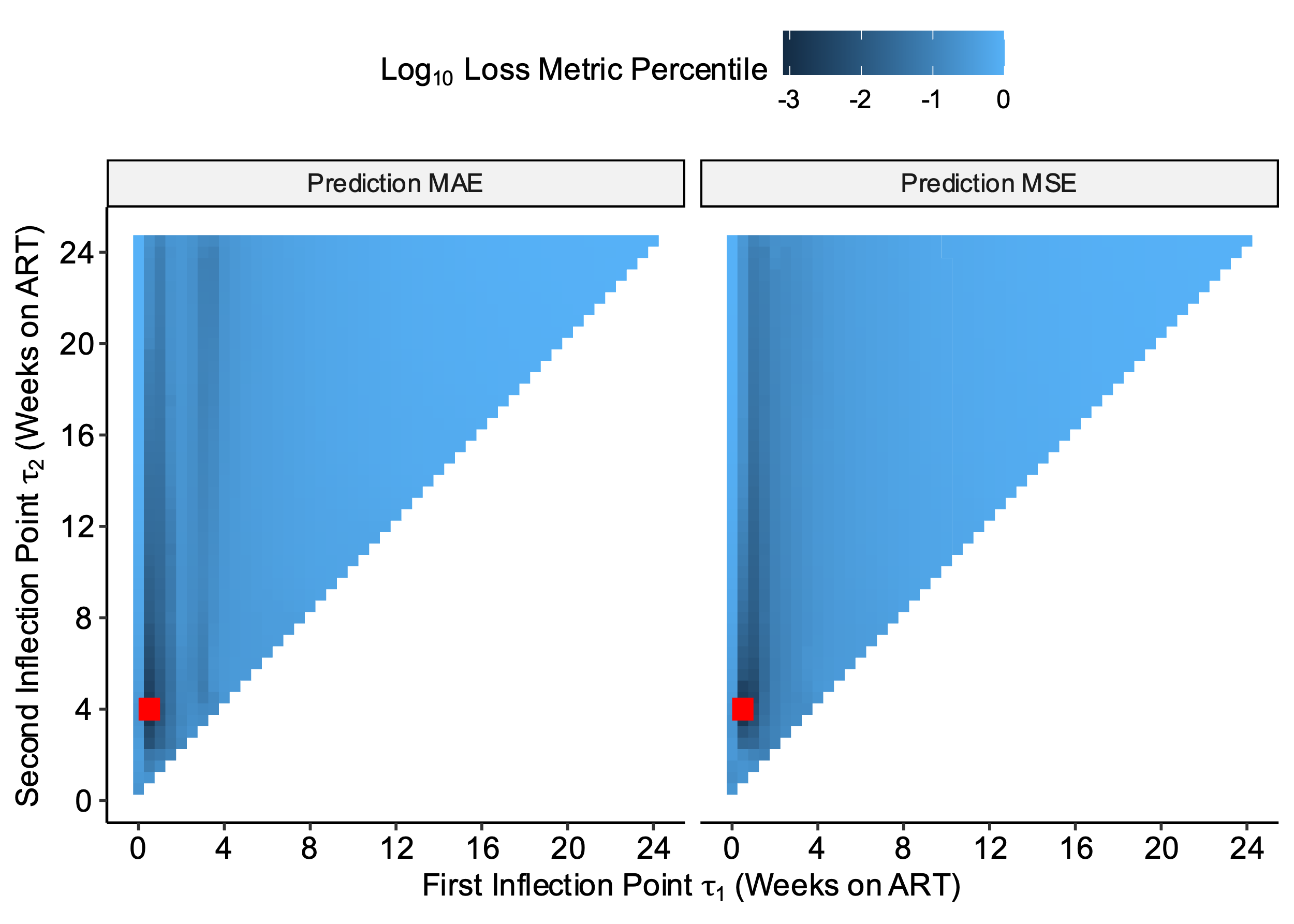

**Supplementary Table 5. Model estimates of plasma HIV RNA decay rates during acute treated HIV.** Slope and half-life (t1/2) estimates of plasma HIV RNA decay rates for models adjusted for initial CD4+ T cell count and timing of ART initiation during frequently sampled timepoints (weeks 0-24). Models included a random intercept for each participant. We calculated estimates for each phase of the final triphasic decay model: 0 to 0.5 weeks, 0.5-4 weeks, and 4-24 weeks. We did not observe a statistically significant decay of plasma HIV RNA after week 4, given that the majority of participants had undetectable plasma HIV RNA levels after a median of 4.14 weeks on ART.

| **Time on ART** | **N** | **Estimate** | **SE** | **P** | **Decay Rate per Day (%)** | **Half Life (Days)** | **Half Life (lower CI)** | **Half Life**  **(upper CI)** |
| --- | --- | --- | --- | --- | --- | --- | --- | --- |
| **Phase 1: 0 to 0.5 weeks** | 67 | -10.6 | 0.974 | 1.86e-24 | 65.0 | 0.659 | 0.541 | 0.778 |
| **Phase 2: 0.5 to 4 weeks** | 67 | -1.42 | 0.140 | 1.14e-21 | 62.6 | 4.93 | 3.98 | 5.89 |
| **Phase 3: 4 to 24 weeks** | 67 | -0.00650 | 0.0166 | 6.95e-01 | 0.449 | 1080 | -4310 | 6460 |

**Supplementary Fig. 12: Predicted versus observed plots demonstrating model performance for plasma HIV RNA**. Validation for the final model for plasma HIV RNA decay was initially performed by looking at the plots of predicted vs observed plasma HIV RNA counts. This plot shows that the triphasic model produces unbiased estimates for a large range of plasma HIV RNA counts. The model systematically underestimates very large plasma HIV RNA values and has high variability at very low plasma HIV RNA values (e.g., <40 copies/mL). Red dashed line denotes the idealized fit where predicted values exactly equal observed values.

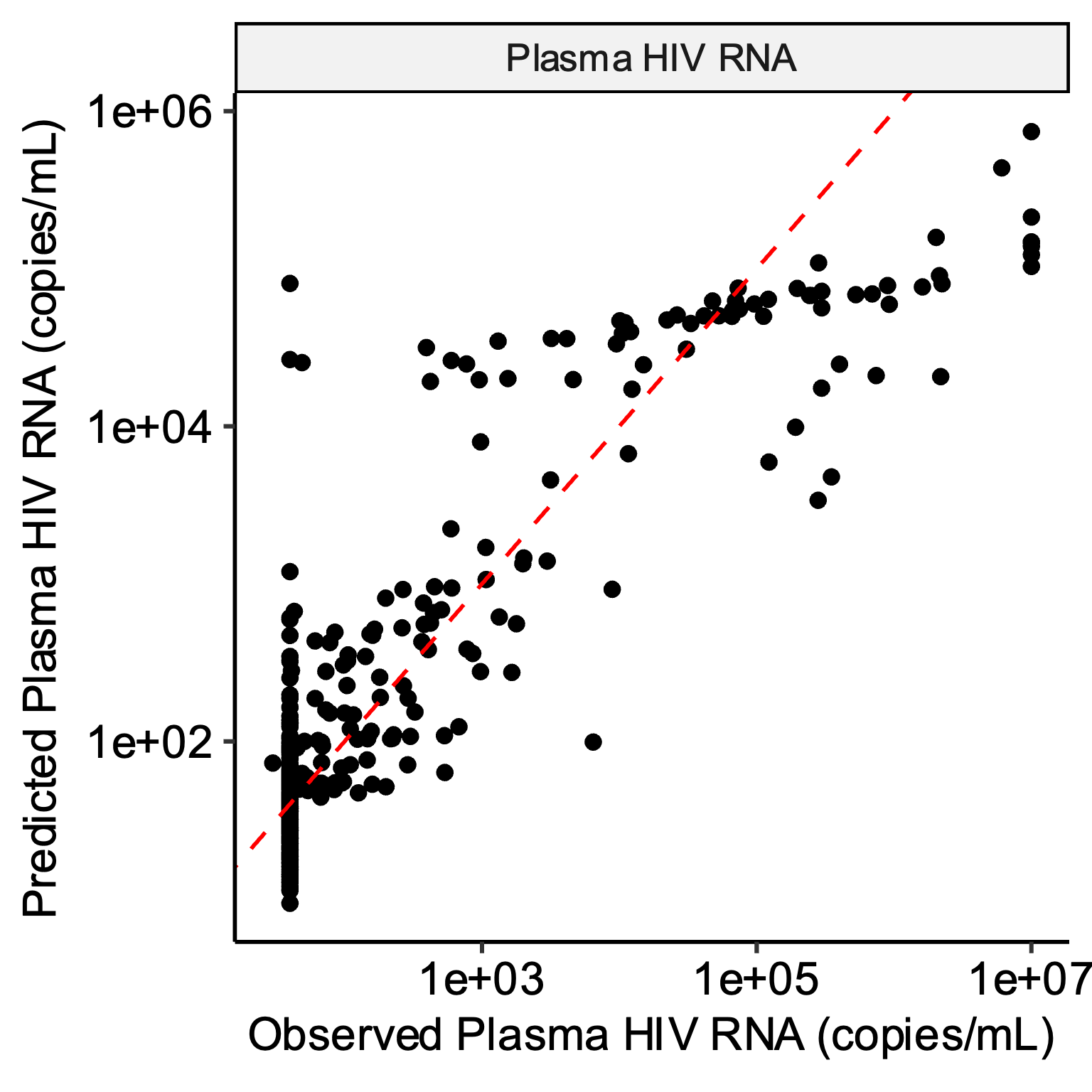

**Supplementary Fig. 13: Plasma HIV RNA decay patterns were associated with known clinical factors associated with HIV reservoir size**. The observed plasma HIV RNA data are shown as thin grey lines for each participant. Plasma HIV RNA decay was faster among participants initiating ART earlier (<30 days vs. 30-100 days) (a) and with higher initial CD4+ T cell counts (shown by tertiles) (b).

**a.**

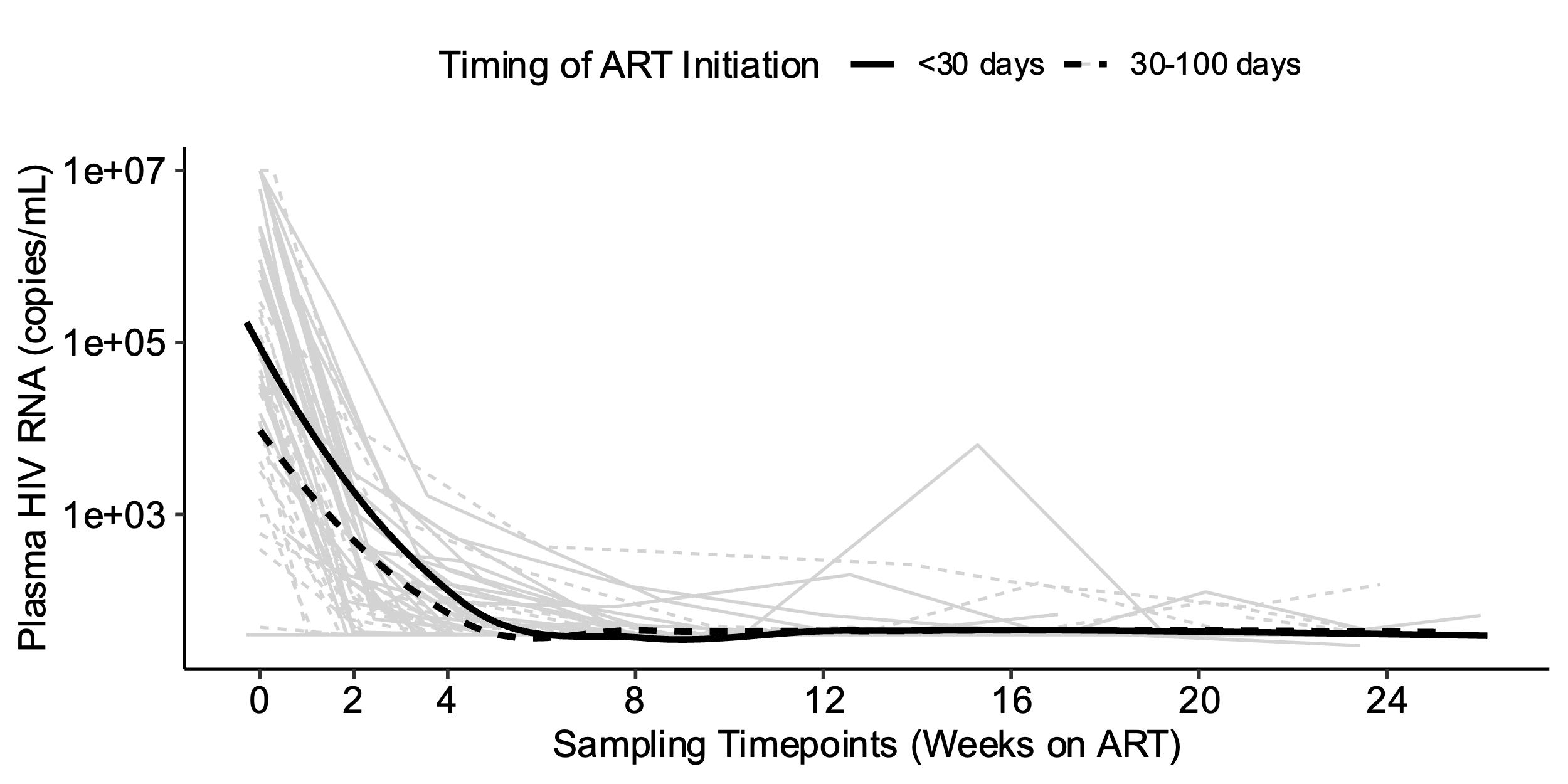

**b.**

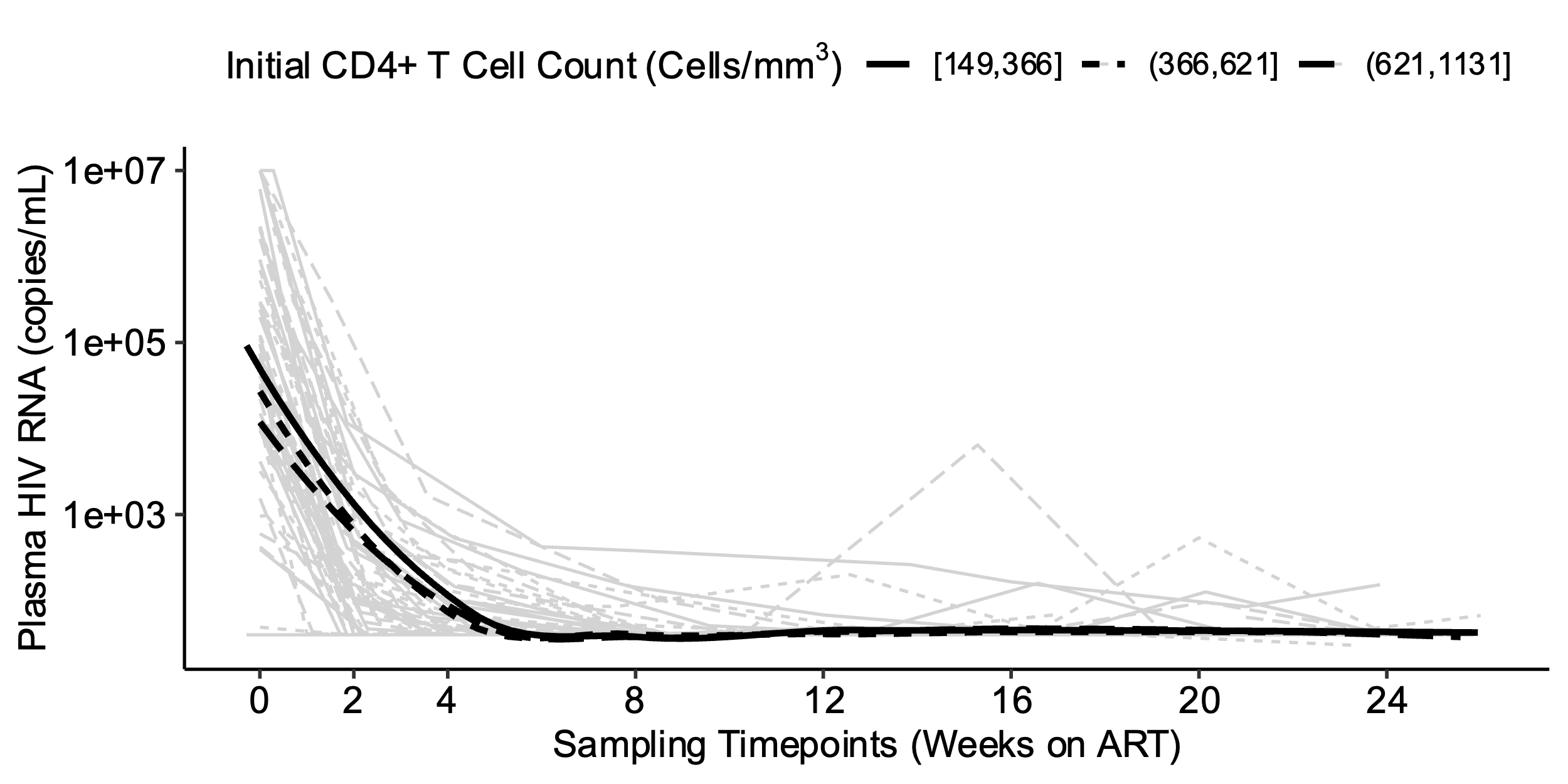

**Supplementary Fig. 14: Predicted plasma HIV RNA decay rates, by tertiles of clinical factors associated with HIV reservoir size.** We performed bootstrapping to estimate the average predicted decay rates of plasma HIV RNA, stratified by known clinical factors associated with HIV reservoir size: timing of ART initiation (a) and initial CD4+ T cell count (b).

**a.**

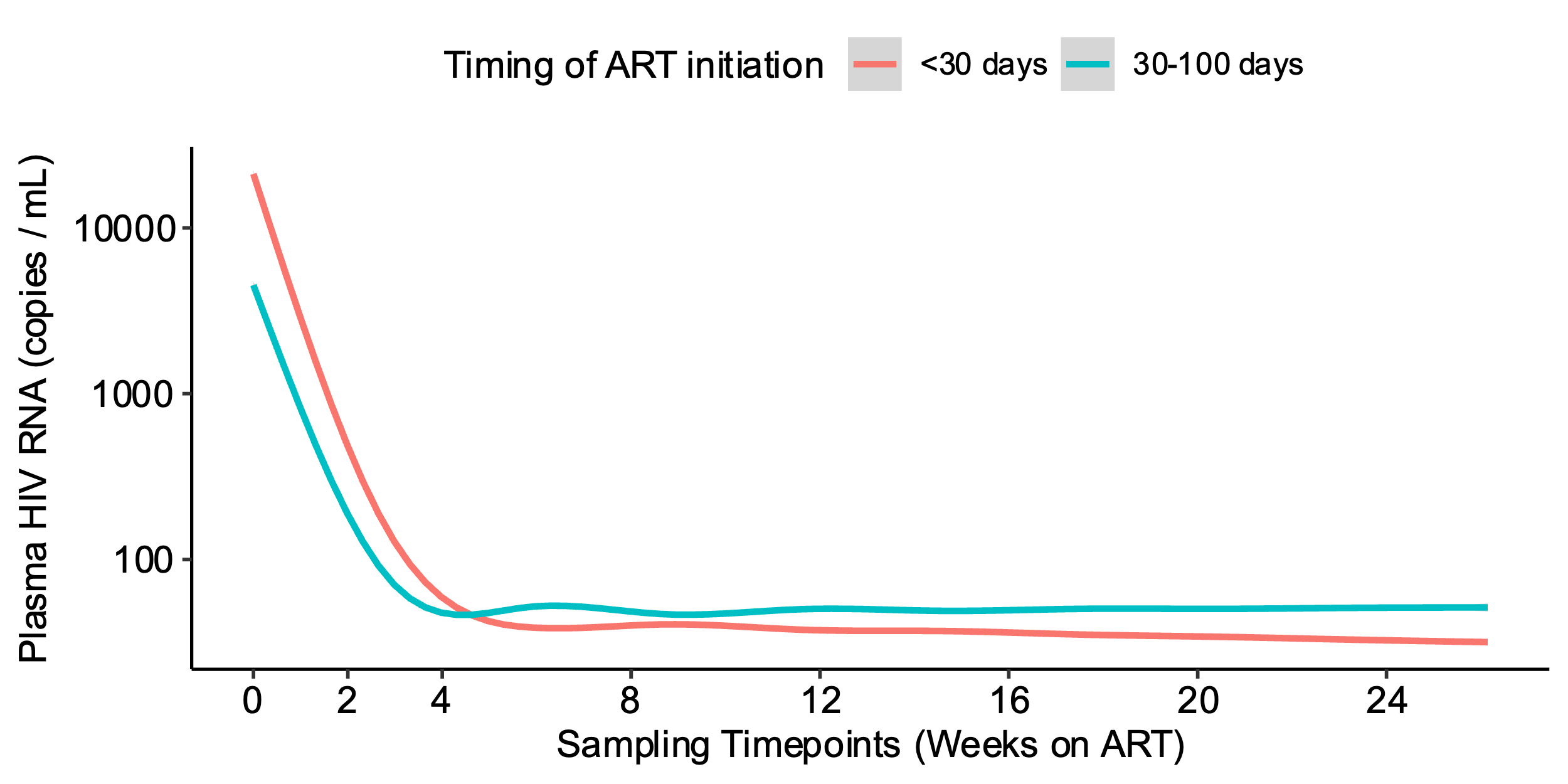

**b.**

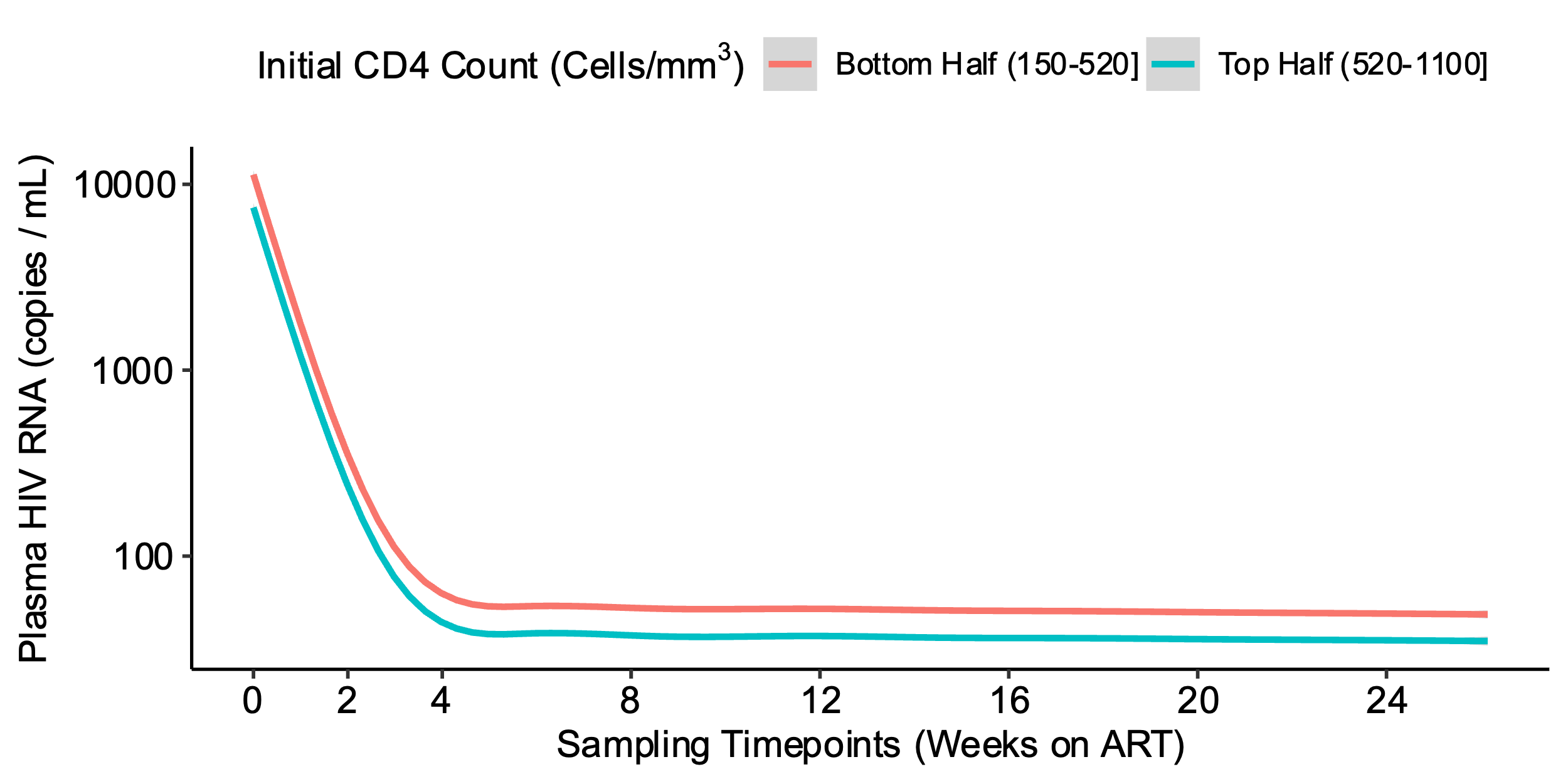

**Supplementary Fig. 15: Fitted spline models to estimate the effect of higher initial CD4+ T cell count on plasma HIV RNA decay rate.** Fitted spline model (red line) with corresponding 95% confidence interval (blue dashed line) is shown for plasma HIV RNA in relation to initial CD4+ T cell count.

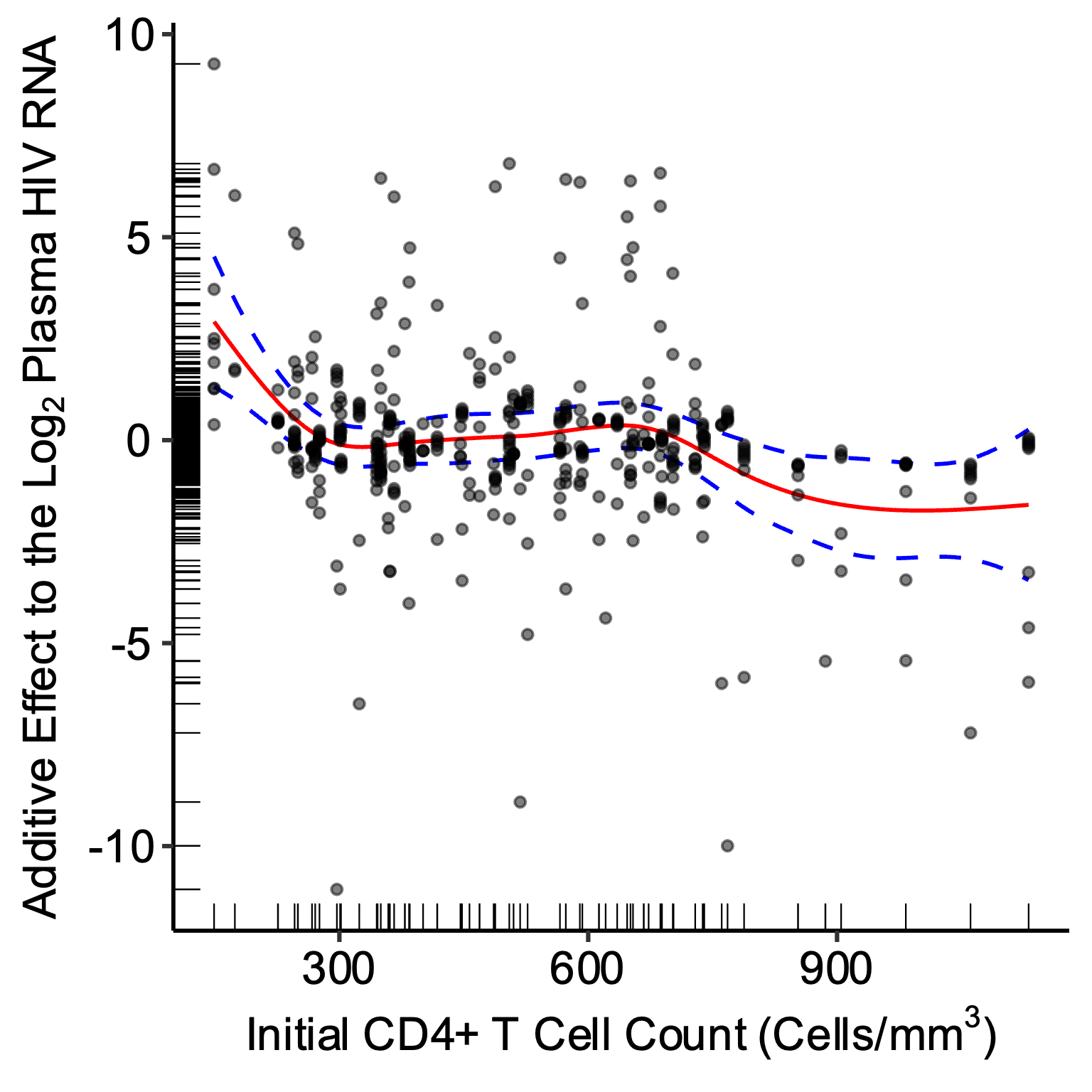
